# A User Preference Analysis Of Commercial Breath Ketone Sensors to Inform the Development of Portable Breath Ketone Sensors for Diabetes Management in Young People

**DOI:** 10.1101/2021.10.29.21265562

**Authors:** Nicola Brew-Sam, Jane Desborough, Anne Parkinson, Krishnan Murugappan, Eleni Daskalaki, Ellen Brown, Harry Ebbeck, Lachlan Pedley, Kristal Hannon, Karen Brown, Elizabeth Pedley, Genevieve Ebbeck, Antonio Tricoli, Hanna Suominen, Christopher J. Nolan, Christine Phillips

## Abstract

**Background:** Portable breath ketone sensors may help people with Type 1 Diabetes Mellitus (T1DM) avoid episodes of diabetic ketoacidosis; however the design features preferred by users have not been studied. We aimed to elucidate breath analysis and design preferences associated with commercial breath ketone devices among young people with T1DM aged 12-16 years and their parents in order to inform the development of a breath ketone sensor prototype for diabetes management.

**Research Designs and Methods:** Two commercially available breath ketone sensors, designed for ketogenic diet monitoring, were explored over one week by ten young people with T1DM to get an insight into breath measurements. Participants interacted with the devices at least twice daily for five days, taking breath ketone, blood ketone and blood glucose measurements. Semi-structured interviews were conducted post-testing with the young participants and their caregivers to elicit preferences related to breath analysis and to inform the co-design of a diabetes breath ketone sensor prototype. To validate the results from a professional healthcare perspective, we interviewed two diabetes educators working in pediatric care about their perspective of young people using breath ketone sensors.

**Results:** Participants acknowledged the non-invasiveness of breath sensors as compared to blood testing. Affordability, reliability and accuracy were identified as prerequisites for breath ketone sensors used for diabetes management. Design features valued by young people included portability, ease of use, sustainability, readability and suitability for use in public. The time required to use breath sensors was similar to that for blood testing. The requirement to maintain a 10-second breath exhalation posed a challenge for users. Diabetes educators highlighted the ease of use of breath devices especially for young people with insufficient blood ketone testing.

**Conclusions:** Breath ketone sensors for diabetes management bear potential to facilitate ketone testing in young people. Our study affirms features for young people that drive usability of breath sensors among this population, and provides a model of user preference assessment.

**Highlights/new findings:**

- Portable breath ketone sensors potentially offer a non-invasive alternative to blood ketone testing in diabetes management, especially for young people with diabetes.
- A user preference assessment of portable non-medical breath ketone devices among young people with Type 1 Diabetes Mellitus demonstrates the key values associated with breath ketone devices for diabetes management.
- The time to test for breath ketones was similar to that of blood ketone testing.
- Young people can and should be engaged in co-creation of medical devices that they will use.

## Introduction

Managing Type 1 Diabetes Mellitus (T1DM) during childhood and adolescence is challenging for young people, their families and caregivers^1 2^. Diabetic ketoacidosis (DKA) is a common complication of T1DM, with an annual risk of 1-10%^3 4^. DKA is a medical emergency with a high fatality rate if not treated early^5^. This is why persons with T1DM are advised to monitor their blood ketone levels if unwell or if they have elevated blood glucose levels.

Ketosis develops as a consequence of food deprivation due to fasting (mild) such as carbohydrate restriction in ketogenic diets or in response to endurance exercise, or due to starvation (more severe)^6^. Due to low availability of carbohydrate as an energy source, fatty acids are released from adipose stores as an alternate energy supply. The liver produces ketone bodies from fatty acids, including β-hydroxybutyrate and acetoacetate, and acetone which is released into breath^6 7^. In T1DM, in which severe deficiencies of insulin can occur, this process of lipolysis and ketogenesis can be uncontrolled resulting in DKA, usually in association with severe hyperglycemia^5 7^.

Studies have found a significant relationship between the blood ketone body ß-hydroxybutyrate and breath acetone exhaled during respiration, including in response to ketogenic diets and during episodes of DKA^7-10^. Thus, the measurement of the breath acetone concentration provides a non-invasive method to measure the level of ketosis^8^ (**Table 1**), and if developed and validated for use in T1DM could help in prevention of severe DKA through early detection. Non-invasive technologies are of particular interest for young patient groups, with studies reporting more fear and pain related to finger pricking and injections among younger children^11^.

**Table 1.** Breath acetone concentrations as reported in Anderson, 2015^8^

|  |  |
| --- | --- |
| Healthy individuals | 0.5 to 2.0 ppm |
| Adults on ketogenic diets (e.g., high fat with low carbohydrate) | elevated levels of up to ~40 ppm |
| Children with epilepsy on ketogenic diets | as high as 360 ppm |
| Fasting | up to ~170 ppm |
| Poorly controlled diabetes can cause ketoacidosis | up to 1250 ppm |

*Notes.* ppm = parts per million

Development of sensing technologies to detect breath acetone for diabetes management is underway^12^, but there are no portable breath ketone sensors that are marketed for diabetes yet. Currently, commercial breath ketone sensors designed for purposes such as monitoring a ketogenic diet for weight loss^8^ or endurance performance^13^ are available. Breath acetone has been described as a good indicator of ketosis in people on ketogenic diets^14^. Breath ketone sensors developed to manage a ketogenic diet can serve as a starting point to understand the value and design of breath ketone sensors for diabetes management.

To date, there are no published studies about user preferences regarding diabetes-specific breath ketone sensors for young people with T1DM, even though user preference and usability evaluation is typically applied in the early design of technology^15^. We aimed to elucidate breath analysis and design preferences associated with breath ketone sensors from the perspectives of young people with T1DM aged 12-16 years and their caregivers, to inform the development of a co-designed breath ketone sensor prototype for diabetes management. For result validation, we added a professional perspective on breath ketone sensor use in young people with T1DM.

## Methods

### Study design

We employed a user preference analysis with young people living with T1DM and their caregivers. User preference analysis has previously been used with diverse (qualitative and quantitative) methods and on various diabetes technologies^16 17^. We chose a qualitative approach based on semi-structured user interviews. With no breath ketone sensor for diabetes management on the market, we used commercially available breath ketone sensors as a starting point in order to inform the development of a diabetes-specific breath ketone sensor. Our young study participants living with diabetes reported no previous experiences with breath devices for diabetes management. Thus, we followed an approach similar to a usability testing method^18 19^ but with an ultimate focus on user preferences. Similar to usability testing procedures^18^, (a) we delivered an explanation of the procedure and instructions for invited participants before (b) these interacted with the commercial breath ketone devices in order to explore breath ketone measurements in contrast to blood measurements, as well as features and design of breath ketone devices. In the next step (c), we collected breath and blood data, notes and comments from the participants regarding the device interaction, and finally (d) assessed the user experience and user preferences in semi-structured online or telephone interviews. In contrast to standard usability testing procedures, we did not use observation during the user-device interaction as our focus was mainly on user preference outcomes (rather than usability), which were assessed using interviews after completion of the device interaction/testing period.

### Pre-selection of commercial devices and development of study protocols

Four different commercially available breath ketone sensors used to assist people on a ketogenic diet were examined: House of Keto Monitor^20^, Ketonix^21^, KetoPro KHC M3 by Keto Health Care^22^, and Keyto Breath Sensor^23^ (**Table 2**).

**Table 2.** Comparison of Breath Ketone Sensors for Ketogenic Diet

| Breath device | Version (year) | Scale/level information | Source |
| --- | --- | --- | --- |
| House of Keto Monitor<sup>20</sup> | House of Keto Monitor (2020) | Results measured in g/l. “The highest documented is 1.9 g/l”, “anything above 0.2 g/l on the device indicates nutritional ketosis state”. | Contact with company |
| Ketonix<sup>21</sup> | Ketonix Basic with USB cable (2020) | Results measured in ppm, “the Ketonix breath ketone range is up to 250 PPM” | Contact with company |
| KetoPro KHC M3 by Keto Health Care<sup>22</sup> | KHC M3 breath meter/KetoPro (2020) | “The KHC M3 breath meter uses mg/l as type of measurement to detect acetones in the breath”<sup>27</sup> | Website |
| Keyto Breath Sensor<sup>23†‡</sup> | Keyto Breath Sensor (2020) | 0 ppm = Levels 1-2 = Not in ketosis; 1-4 ppm = Levels 3-4 = Light/almost in ketosis (0,01- 0,04 mmol/l); 5-9 ppm = Level 5-6 = Ketosis (0,05- 0,09 mmol/l); >10 ppm = Level 7-8 = High ketosis (>0,1 mmol/l) ‡<br>“We chose not to report acetone concentrations in PPM or to attempt to convert PPM to blood β-hydroxybutyrate (mmol/L). The Keyto Level system was simply more effective, motivating, and fun without adding complexity and false precision”<sup>28</sup> | Contact with company and "Keyto Fundamentals" (instructions) |

Notes. 1ppm = approx. 1mg/l; Only Ketonix was registered at FDA (US) and Läkemedelsverket (SE) as a Non-Invasive Class I Medical Device<sup>29</sup> at the time of data collection, while the other devices were not FDA-approved.
† This Keyto Breath Sensor version required an app for result display
‡ It was unclear if higher ppm levels could be measured as well, the manufacturer did not provide clear information

Four young people with T1DM who are part of our research team in our consumer-focused research program (two male and two female) pre-tested the sensors at home for two to three weeks and provided detailed oral and written feedback in discussions. Based on their feedback, KetoPro and House of Keto Monitor sensors were assessed to be most suitable for further testing with study participants. Keyto and Ketonix devices were excluded because Keyto required an app to operate the device, increasing complexity for use. The study focus was on breath ketone sensors rather than on additional connected devices. Ketonix did not report numerical readings. Device testing instructions (**Appendix 1**) and a structured interview protocol (**Appendix 2**) were developed based on the pre-testing of the devices and the feedback provided by our young research team members, our clinical team members (endocrinologists), our previous studies about experiences with diabetes technologies^24 25^, and breath sensor literature^12 26^. The focus of the interview guide was both on actual functioning and potential functioning of the sensors (improvements and potential additional features). The whole research team refined the instructions and interview guide continuously in the process of study preparation.

### Recruitment

Ten young people with T1DM who attended the Endocrinology Department of Canberra Hospital and had participated in a previous study with our research team, were invited, and agreed, to test one of the two selected sensors at their homes (KetoPro n=4; House of Keto Monitor n=6). As a result of the COVID-19 pandemic, all studies were conducted in a contactless manner.

### Data collection and analysis

The breath ketone sensors were posted to each of the ten participants in June/July 2020, together with test strips for five days of blood ketone and blood glucose measurements (twice daily). Following the four described steps^18^, device testing instructions (**Appendix 1**) were provided through a telephone or video call and in written form together with a spreadsheet to record testing results. During the five-day testing period, participants interacted with the devices at least twice daily. Breath ketone, blood ketone and blood glucose measures were taken in parallel before breakfast and after school but before eating, as ketone readings were expected to be higher after a period of fasting. We did not aim to conclude about the commercial devices’ (in)accuracy with breath and blood measurements taken throughout the testing period. Because testing did not happen in a clinical setting (testing for DKA) and because our participants were mostly not sick (no risk of DKA), and were not on a ketogenic diet, low (or even no) ketone readings were to be expected. The purpose of the breath-blood testing was not to evaluate device accuracy, but to trigger feedback about breath device usage in contrast to blood testing. Following the user-device interaction, online video- or telephone-interviews were conducted with participants accompanied by their parent(s) to elucidate their breath device preferences based on the experiences with the use of the commercial breath ketone devices. The use of the commercial devices gave the young people a sense about what a breath sensor device can deliver and how it works. In this way, the use of the commercial devices triggered feedback which was valuable for informing the development of a breath device specifically designed for diabetes management.

In order to validate the feedback from young people and their caregivers from a professional healthcare perspective and to set the user input in relation to a healthcare perspective, two online video interviews with diabetes educators working in pediatric care were conducted in addition. The professional perspective on young people using breath ketone sensors helped to comprehend young people’s feedback in more detail, and added important details and background information on breath sensor testing in youth. A separate short semi-structured interview protocol was used for the interviews with the diabetes educators (**Appendix 3**). It focused on information about ketone testing in young patients from a healthcare professional perspective, previous experiences with breath analysis, and expectations associated with breath sensors for the use by young people with T1DM.

The summary of findings about preferences and ideal features as described by the participants enabled the compilation of a list of preferred breath sensor characteristics to inform the development of co-designed breath ketone sensor prototypes for diabetes management. Ethical approval for the study was obtained by the Australian National University’s Human Ethics Committee and ACT Health Human Research Ethics Committee (HREC, Australia) in April/May 2020 (2019.ETH.00143; 2019/ETH12170). The study participation was based on informed consent (written, signed consent form). For minors, written consent was signed by a parent/guardian.

## Results

### Sample description

All participants (**Table 3**) finished the minimum of five days breath and blood testing twice per day and participated in online video or telephone interviews. One person only provided four days’ test results, despite participating for five days.

**Table 3.** Participant characteristics and devices tested

| No | Device | Gender | Year of diagnosis (range) | CGM | Insulin pump | Other technology |
| --- | --- | --- | --- | --- | --- | --- |
| 1 | KetoPro KHC M3 | M | 2011-2015 | Medtronic/ Guardian | Medtronic 640G |  |
| 2 | KetoPro KHC M3 | F | 2006-2010 | Dexcom G5 | Medtronic |  |
| 3 | KetoPro KHC M3 | F | 2011-2015 | Dexcom | Tandem T: Slim |  |
| 4 | KetoPro KHC M3 | M | 2006-2010 | Dexcom | Medtronic |  |
| 5 | House of Keto Monitor | F | 2016-2020 | Dexcom | Tandem T: Slim |  |
| 6 | House of Keto Monitor | M | 2006-2010 | Dexcom | Medtronic 640G |  |
| 7 | House of Keto Monitor | M | 2011-2015 | (previously Dexcom G5) | Medtronic | Free-style Libre (temporary) |
| 8 | House of Keto Monitor | M | 2016-2020 | Dexcom | Tandem T: Slim | Free-style Libre |
| 9 | House of Keto Monitor | M | 2006-2010 | Medtronic/ Guardian | Medtronic 670G |  |
| 10 | House of Keto Monitor | F | 2016-2020 | Dexcom G5 | --- |  |

Note. Age range 12-16 years

### Baseline data

No participant had previously used a breath ketone sensor or any other breath device for their diabetes management. Reasons cited by participants included not being aware that breath ketone measurements were possible (participants #2, 5-7, 9, 10; data excerpts in **Table 4**), that there was no opportunity or no device available prior to our study (#3, 4, 6), or that there was no perceived need for breath analysis (#8). All participants said they had previously used blood ketone tests when they were either sick (#1-10), had very high blood glucose over a period of time such as extended values over 15mmol/L (#1, 3-7, 10), or did not know why the blood glucose was high, or why the insulin was not effective (#1). One participant mentioned blood ketone testing as a “*backup*” option (#1). Overall, blood ketone testing use was “*very infrequent*” (#7) and only in particular circumstances such as those described above, and was not monitored regularly as compared to blood glucose testing.

**Table 4.**
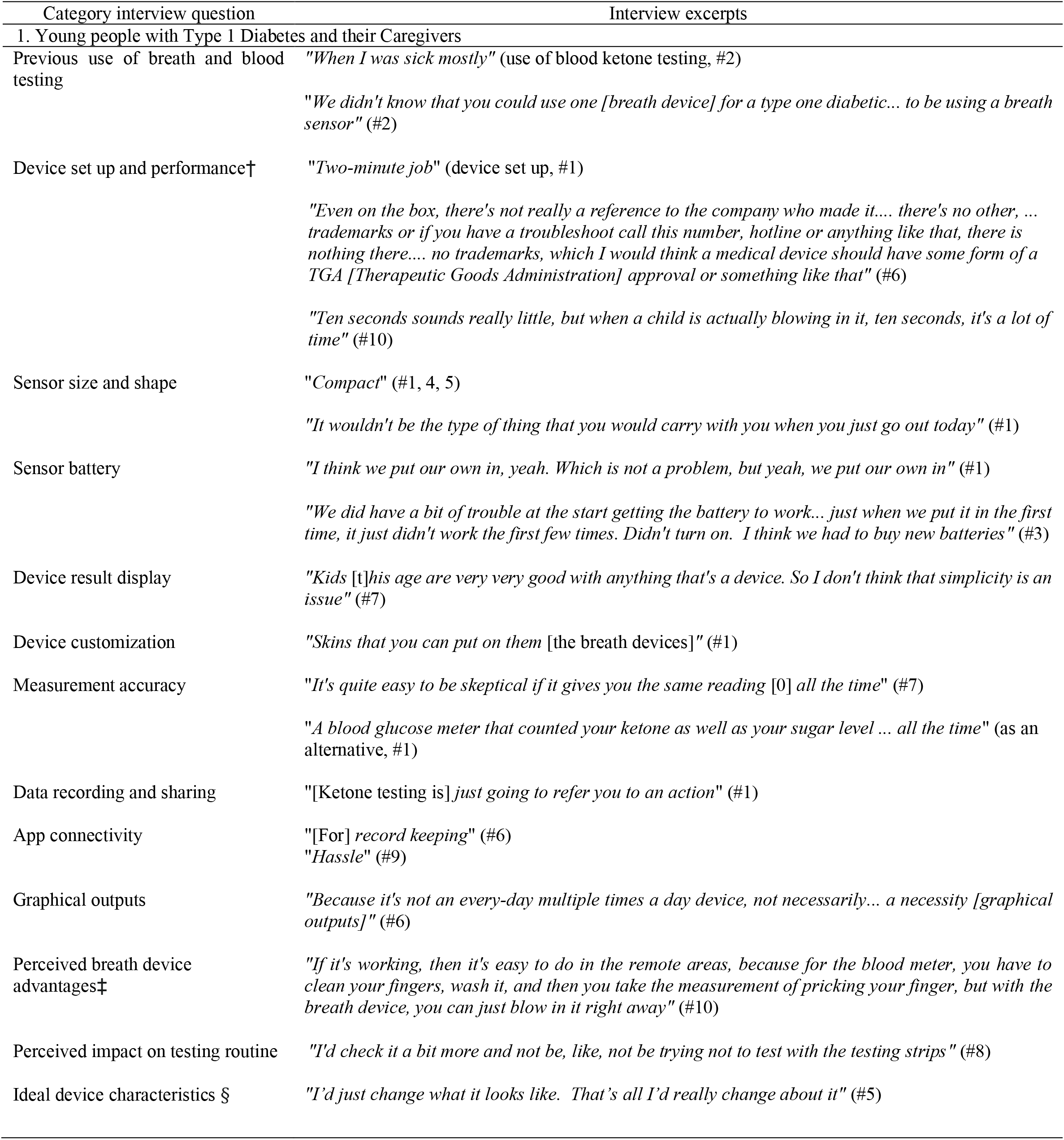
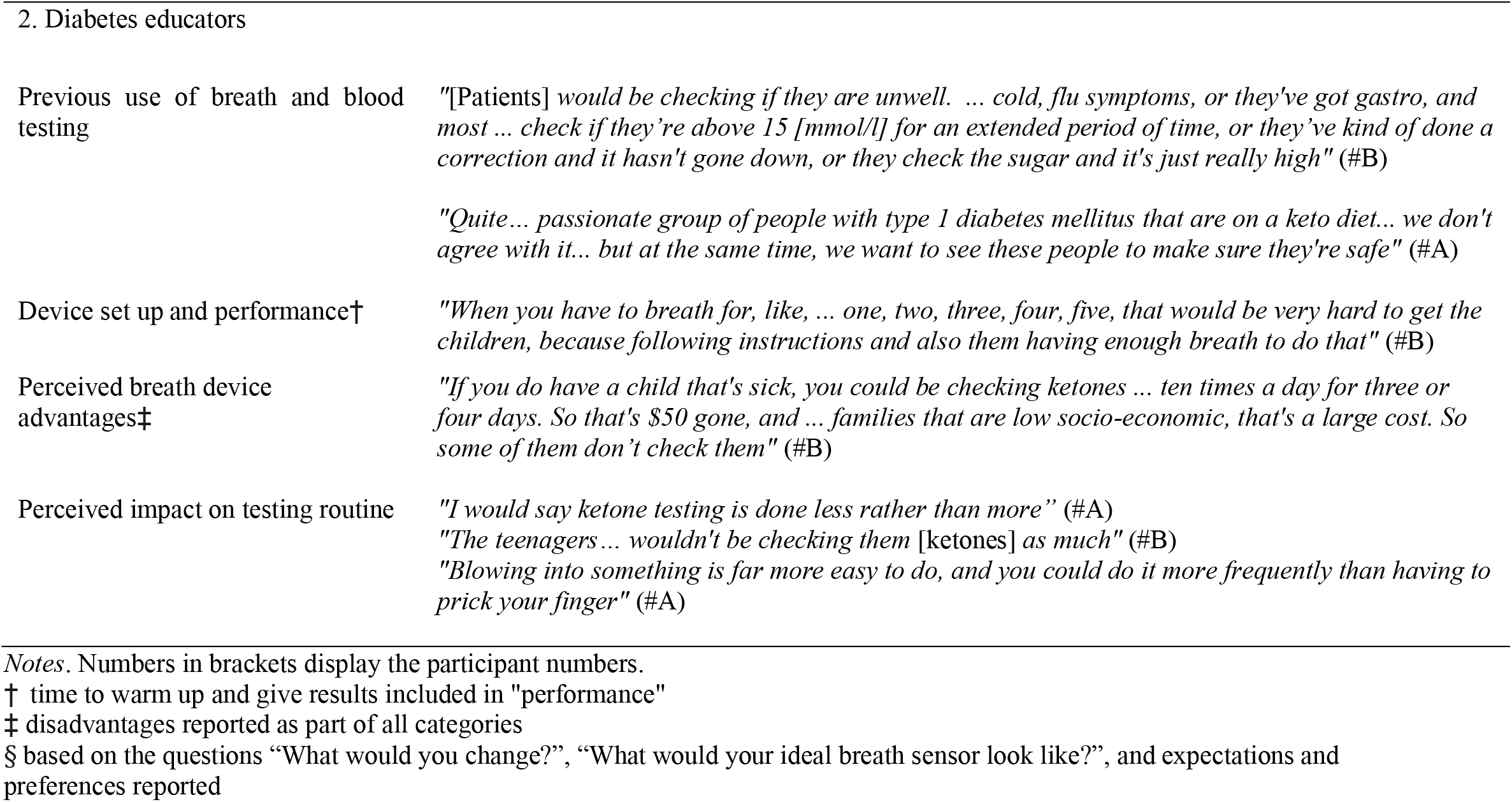
Interview excerpt examples

| Category interview question | Interview excerpts |
| --- | --- |
| 1. Young people with Type 1 Diabetes and their Caregivers |  |
| Previous use of breath and blood testing | "When I was sick mostly" (use of blood ketone testing, #2)<br>"We didn't know that you could use one [breath device] for a type one diabetic... to be using a breath sensor" (#2) |
| Device set up and performance† | "Two-minute job" (device set up, #1)<br>"Even on the box, there's not really a reference to the company who made it.... there's no other, ... trademarks or if you have a troubleshoot call this number, hotline or anything like that, there is nothing there.... no trademarks, which I would think a medical device should have some form of a TGA [Therapeutic Goods Administration] approval or something like that" (#6)<br>"Ten seconds sounds really little, but when a child is actually blowing in it, ten seconds, it's a lot of time" (#10) |
| Sensor size and shape | "Compact" (#1, 4, 5)<br>"It wouldn't be the type of thing that you would carry with you when you just go out today" (#1) |
| Sensor battery | "I think we put our own in, yeah. Which is not a problem, but yeah, we put our own in" (#1)<br>"We did have a bit of trouble at the start getting the battery to work... just when we put it in the first time, it just didn't work the first few times. Didn't turn on. I think we had to buy new batteries" (#3) |
| Device result display | "Kids [t]his age are very very good with anything that's a device. So I don't think that simplicity is an issue" (#7) |
| Device customization | "Skins that you can put on them [the breath devices]" (#1) |
| Measurement accuracy | "It's quite easy to be skeptical if it gives you the same reading [0] all the time" (#7)<br>"A blood glucose meter that counted your ketone as well as your sugar level ... all the time" (as an alternative, #1) |
| Data recording and sharing | "[Ketone testing is] just going to refer you to an action" (#1) |
| App connectivity | "[For] record keeping" (#6)<br>"Hassle" (#9) |
| Graphical outputs | "Because it's not an every-day multiple times a day device, not necessarily... a necessity [graphical outputs]" (#6) |
| Perceived breath device advantages‡ | "If it's working, then it's easy to do in the remote areas, because for the blood meter, you have to clean your fingers, wash it, and then you take the measurement of pricking your finger, but with the breath device, you can just blow in it right away" (#10) |
| Perceived impact on testing routine | "I'd check it a bit more and not be, like, not be trying not to test with the testing strips" (#8) |
| Ideal device characteristics § | "I'd just change what it looks like. That's all I'd really change about it" (#5) |
| 2. Diabetes educators |  |
| Previous use of breath and blood testing | *"[Patients] would be checking if they are unwell. ... cold, flu symptoms, or they've got gastro, and most ... check if they're above 15 [mmol/l] for an extended period of time, or they've kind of done a correction and it hasn't gone down, or they check the sugar and it's just really high" (#B)*<br>*"Quite ... passionate group of people with type 1 diabetes mellitus that are on a keto diet... we don't agree with it... but at the same time, we want to see these people to make sure they're safe" (#A)* |
| Device set up and performance† | *"When you have to breath for, like, ... one, two, three, four, five, that would be very hard to get the children, because following instructions and also them having enough breath to do that" (#B)* |
| Perceived breath device advantages‡ | *"If you do have a child that's sick, you could be checking ketones ... ten times a day for three or four days. So that's \$50 gone, and ... families that are low socio-economic, that's a large cost. So some of them don't check them" (#B)* |
| Perceived impact on testing routine | *"I would say ketone testing is done less rather than more" (#A)*<br>*"The teenagers... wouldn't be checking them [ketones] as much" (#B)*<br>*"Blowing into something is far more easy to do, and you could do it more frequently than having to prick your finger" (#A)* |

Notes. Numbers in brackets display the participant numbers.
† time to warm up and give results included in "performance"
‡ disadvantages reported as part of all categories
§ based on the questions "What would you change?", "What would your ideal breath sensor look like?", and expectations and preferences reported

Overall, all participants said that breath ketone sensors must be accurate and reliable (#1-10) and all but three participants (#2, 7, 10) reported they would choose breath over blood if that choice was available and if breath sensors were approved and accurate (#6).

Diabetes educators confirmed the usefulness of the sensors, especially as blood ketone testing by their patients was being followed less than was recommended, impacted by the price and availability of ketone strips. Breath ketone sensors were expected to be *“very valuable”* (#A) for insulin pump users as pump users *“can develop ketones far more quickly*” (#A) compared to individuals on multiple daily injection regimens with *“long-acting insulin sitting in the background”* (#A). One general challenge mentioned by the nurse educators (participants #A and B) related to people with T1DM who choose to follow a ketogenic diet. Such a diet generally was reported to impact ketone readings and was perceived as unsafe for youth with T1DM, thus this group required particular attention (#A).

### Measurement accuracy

Despite participants expectation of low ketone readings (#4, 7), accuracy remained a concern, and was reported to be the major obstacle to breath sensor use (#7). Breath readings frequently displayed no results as shown in **Appendix 4**. Some participants questioned if the breath ketone sensor was working, especially when a zero (0) reading was repeatedly shown after breath tests (#3, 5-7, 9, 10) (**Appendix 4**). Participants were unsure about device reliability (#2, 9) and did not “*feel like you can trust it*” (#7). One young person wondered if toothpaste or other substances that included alcohol such as mouth wash might affect readings (#1). Another participant questioned whether the way of breathing into the device, such as interrupted breathing, affected device readings (#9). Breath sensor accuracy concerns (#7, 10) led some participants to favour blood tests over breath tests, and two said they would either prefer blood tests (#2), or wanted to avoid additional devices as the blood meter measured both ketone and glucose levels (#10). Accuracy was confirmed to be one of the most crucial preconditions for the breath sensors by the diabetes educators (#A and B).

### Breath ketone device set up and performance

Device set-up was considered to be easy and fast to complete (#1). Instructions to use the breath ketone sensors were described as “*quite basic*” (#2), and only available in English language (#1). The instructions did not deliver information about how to buy extra mouthpieces (#6) or how often to change mouthpieces for the House of Keto Monitor (#6, 9), while the KetoPro recommended mouthpieces be changed after 10-15 uses to avoid incorrect readings (#1). Some participants washed or sanitized mouthpieces for reuse (#1, 5, 7, 9, 10), two people used only one mouthpiece throughout (#3, 8), while one felt uncomfortable reusing them (#2). KetoPro provided six initial mouthpieces, while House of Keto Monitor provided four, which was felt to be too few by one participant (#9). Mouthpieces kept falling off in one case (#8). Overall, clear information was missing about how to use mouthpieces (#5, 6), or how to breathe into the devices (#9).

Information about the device manufacturers was considered insufficient by some participants, with no clear provision of company website addresses (#6). One participant suggested that a breath ketone sensor for diabetes management would need a clear indication that it is a medical device (#3). Trademarks or TGA (Therapeutic Goods Administration) approval were perceived as important for diabetes breath sensors (#6).

The devices showed results immediately after testing (#7). Only one participant had problems with incorrect signals, as the light that indicated the sensor was ready for use did not work properly (#3). In general, it was estimated that a breath test took approximately the same time as a blood test (#7, 9). Device warm-up time was reported to be 10-20 seconds, with 20 seconds being perceived as too long by one participant (#3). More concerning was that the devices required a breath of 10 seconds to achieve a test result, which was challenging for six participants (#2-4, 8-10). One participant suggested that the user should not have to actively breathe into the breath sensor but just count out loud close to the device similar to advanced alcohol testing devices (#9).

Similar to the young participants, one diabetes educator highlighted potential breathing difficulties in children with a breath device (#B).

### Sensor size and shape

Young people did not anticipate carrying breath ketone sensors all the time (#1); one participant described carrying them as “*awkward*” (#4). One mother mentioned that her son had one blood ketone meter stored at home and one at school to avoid having to carry the device with them (#9). The breath sensors could fit into the diabetes pencil case carrier (#9) due to their “*compact*” (#1, 4, 5) design. Half of the participants suggested *“a little bit smaller”* device size (#10, similar 1, 3, 6, 8); yet three participants pointed out that it might be difficult to hold the device if it was too small (#1, 2), especially if one had *“big hands”* (#7). Breath sensor shapes were perceived as comfortable (#3), like *“a good handle”* shape (#1) with finger grips (House of Keto Monitor) being perceived as convenient (#6, 7).

A diabetes educator pointed out that the ease of portability of a breath ketone sensor was a critical determinant of use (#A).

### Sensor battery

There were no issues with battery life within the short time span of testing. Both devices required batteries, which were not provided (#1, 3), while subsequent sensor versions provided options for USB charging (e.g., Ketonix).

### Device result display

All participants emphasized the importance of displaying the actual ketone values instead of seeing a level range or a traffic light result (green, orange, red light). Some thought a traffic light display may be a useful addition to evaluate the result quickly (#1, 4-6, 8). One mother said that simplicity was not an issue with most young people being “tech savvy” (#7). The fact that the breath sensor user could not see the screen while breathing, and thus did not know when to stop was considered problematic (#5). Additional sound was suggested to help with the timings (#5). Moreover, readings disappeared too fast from the screen (#9).

### Device customization

Most participants especially liked the idea of being able to customize the breath ketone sensors (#1, 2, 5-8), for example being able to choose its color. One person suggested customized device “*skins*” (#1).

### Data recording and sharing

Opinions diverged regarding data recording and sharing. Some participants thought data recording was not necessary because the ketone testing was “*just going to refer you to an action*” (#1) in a particular moment and not on a continual basis (#5, 9). Most participants thought it would be good to see previous records (#2-4, 6-8, 10), especially to be able to show them to their healthcare professionals (#8). All but two participants (#4, 9) thought that sharing ketone results with the doctor was beneficial in order to provide the doctor with the “*bigger picture*” (#1, 2, 6-8, 10), even if it was just available in the form of a download option (#6). Ketone data sharing with others (e.g., parents) was wished for “*if someone wasn’t home with the child*” (#5), and for assistance of the young person (#7).

### App connectivity

Some participants expressed interest in having the breath sensor connected to an app (#3, 4). An app was perceived as a potential source of information regarding implications of the measured levels (#1), could be used for “*record keeping*” (#6), to give an overview of trends (#4), and to enable checking (#8). Others thought an app was unnecessary (#5, 6, 7) or a “*hassle*” (#9), and preferred to have the history log in the breath sensor device itself (#2). Two participants mentioned that it would be good to have app connectivity as an option as opinions differed in this regard (#7, 9). Overall, the preference was for the breath ketone sensor to be an autonomous device, with the additional option of app connection.

### Graphical outputs

Graphical outputs did not make sense for participants who were only interested in actual values (#2, 9). The fact that ketones were only measured in certain circumstances (#6, 10) rendered moot the benefits of trend mapping or graphical outputs (#1, 5, 6). Others thought graphical outputs might be a good option (#7) for people who test for ketones often (#8).

### Perceived advantages of breath devices

Advantages of the breath sensor over the blood meter were its non-invasiveness, avoiding the pain and messiness of finger-pricking (#1-7, 10). This was seen as especially beneficial for younger children (#7, 9). Moreover, participants mentioned the convenience of the breath sensor (#1, 3-10), the clean procedure (#5, 7, 8), the compact design without additional required parts (#1, 5, 9), and the suitability for use in remote areas (#10) or when traveling when finger pricking is difficult due to hygiene requirements (#9). Participants also highlighted the improved convenience at school with easy, quick and discreet use of the breath sensor (#4-6).

Sustainability of the breath sensors was considered an advantage, with less waste produced such as used test strips (#1, 3, 7). Breath sensor maintenance was perceived as more convenient than blood device maintenance (#6), due to not having to have a personal stash of spare test strips (#4), given the haphazard commercial availability of ketone strips (#4, 10). Ketone strips were costly (#4, 8), and expired after a certain time if unused (#6).

Diabetes educators emphasized that *“a breath ketone device would make life simpler [for patients]”* (#A). Moreover, similar to the young people one educator highlighted that breath devices could solve the problem of pharmacies frequently being *“out of stock”* of blood ketone strips (#B). In addition, *“ongoing cost”* (#B) could be reduced with a compact breath device without additional required parts (#B).

### Perceived impact on testing routine

Participants predicted that they would test for ketones more frequently if they had a breath sensor because it did not require finger pricking and avoided the use of costly ketone strips (#1, 3, 4, 8- 10). Three participants expected that having a breath sensor would not change their current daily routine (#2, 6, 7).

Both diabetes educators expected that a breath testing method would facilitate more frequent testing which was highly appreciated by healthcare professionals (#A and B). They pointed out that ketone testing was partly not performed often enough by young people (#A and B).

To conclude, **Table 5** summarizes preferred characteristics of breath ketone sensors for diabetes management as mentioned by our young participants.

**Table 5.** Summary of Preferred Breath Ketone Sensor Characteristics

| Breath device features | Explanation |
| --- | --- |
| Accuracy/reliability | • accurate results<br>• reliable device<br>• information about food/medication/other chemicals that can affect the readings provided<br>• specific instructions how to breath into the device for accurate results and how the breathing can affect the readings<br>• information about threshold levels<br>• information about correlation between blood and breath results for potential comparison by the user |
| App connectivity | • app connected to breath sensor optional<br>• app easy to find and to download from app store, easy to operate<br>• breath device should still be autonomous<br>• app for record keeping, delivering background information, trends |
| Battery | • battery or USB charging optional<br>• battery provided |
| Company information | • information about manufacturer of device<br>• contact and helpline options |
| Cost/funding | • affordability of breath devices and mouthpieces<br>• funding options<br>• subsidies |
| Customization | • customization options such as colour and shape of device<br>• “skins” for the device purchasable |
| Data recording/sharing | • data storage/recording optional<br>• data sharing options (e.g., with healthcare team) |
| Display and sounds | • numerical readings,<br>• clear result display,<br>• traffic light results in addition to numerical results,<br>• graphical outputs/trends optional for users testing frequently,<br>• clear indication when to start/stop breathing (sound or display),<br>• correct signals,<br>• readings displayed long enough after testing |
| Manual | • provided in several languages,<br>• easy to understand, yet comprehensive,<br>• maintenance instructions |
| Mouthpieces | • information about how to replace mouthpieces, how often to change them, and where to buy additional mouthpieces, reuse of mouthpieces (sanitising requirements),<br>• sufficient amount of mouthpieces provided |
| Performance (time) | • quick display of testing results,<br>• short warm-up time (less than 20 seconds),<br>• required breath less than 10 seconds,<br>• breath test should not be much longer than blood test |
| Set-up | • easy and fast to set up breath sensor,<br>• instructions easy to understand |
| Shape | • optional square or pen shape to choose from,<br>• finger grips |
| Size | • compact design, small enough to fit in toolbox for diabetes devices,<br>• as small as possible, yet not too small to hold comfortably in hand |
| Testing routine | • specific information about breath testing routine and frequency (specific situations explained in which ketone testing should be used) |
| Trademarks/approval | • indication that the breath sensor is a medical device, trademarks, Therapeutic Goods Administration (TGA) approval,<br>• advanced breath testing without active breathing (similar to some alcohol testing devices that are held close to mouth only) |

## Discussion

Our findings highlight the potential, but also specific challenges, of breath ketone sensors for diabetes management. The non-invasiveness of breath sensors is an advantage over other diabetes devices which cause discomfort^24^. With empirical data from a usability evaluation among young people, our study adds to existing commentary which has outlined the challenges associated with the realization of breath sensors in clinical practice^30^, including breath marker selection, breath sampling, sensitivity, stability, and selectivity requirements, portable device integration, user communication, and clinical applicability.

Our results highlight the importance of providing specific information about how to breathe into the devices, as this might affect the readings. Most breath sensors rely on users emptying their lungs thereby allowing measurement of acetone in “late expiratory breath”^31^, which gives a more accurate reading relative to ketones in blood^32^. This breath sampling method is crucial as it allows measurement of alveolar air, which is the portion that takes part in gas exchange with blood in the pulmonary capillaries^33^. Breathing instructions must be specific and manageable. Our results have shown that the required breath length can be challenging especially for young users.

### Implications for breath ketone sensor development

Incorporating the voices of consumers into medical device design is a key feature of all contemporary medical device design pathways, but barriers to doing this openly and productively remain^34^. Van der Panne et al. note that users may not be skilled in the ability to envision innovation, with a cognitive bias to similarity or incremental adaption^35^. Device designers may suffer from insufficient organizational support to engage with users, or experience in adapting user advice to incorporate device designers^36^. This is particularly the case for products that have users who are dissimilar in socio-economic background and group to the designers^37^.

Our study provides a case study in engaging with young people to assess usability of the device at a very early stage in product development. All our participants were 16 years and younger. The initial stage of sensor selection for usability assessment was performed by other young people with TIDM who are fellow researchers in our consumer-focused research program. The 10 young people who tested the devices were familiar with personalized measuring devices, and highly familiar with technological solutions. The existence of a “like” device (non-medical breath ketone devices) provided an opportunity to road test features which are valuable for a diabetes-specific breath sensor for young people with T1DM, and to engage with their own views to emphasize valued elements. It is worth noting that the more tech-enhanced devices (those requiring an app, or converting data into visual codes or trends) were relatively undervalued attributes by young users. They placed more emphasis on functionality (portability, ease of use and readability) and noted that aesthetics (having a customizable appearance) and being unobtrusive were also important. The latter should not be surprising for young people with a condition which declares itself every time they have to give themselves an insulin injection in public.

Our study demonstrates that co-design approaches that actively involve patients and healthcare professionals can be used at all stages of device design and development^24 34^. Our study findings helped refine the initial design of breath ketone sensor prototypes by informing our engineering team colleagues about the preferred device characteristics reported by participants in this study. The involvement of people with lived diabetes experiences as research team members and consultants similar to our procedure is described in a US paper on patient engagement in diabetes care^38^; to our knowledge ours is the first to describe co-design with young people for breath devices.

### Study Limitations and Future Research

By its very nature, this usability study was small, but sufficient to obtain saturation in analysis. This was not an accuracy or reliability study of measurement by breath ketone sensors, and so was unable to deliver conclusions about the extent of breath sensor (in)accuracy in situations of ketosis or the repeatability of results. The selected breath sensors were not designed for diabetes management, varied in units and scales, and did not provide transparent information about threshold levels. A follow-up clinical study is necessary to assess specifically the accuracy of a breath ketone sensor for T1DM and, hence, whether it is suitable for ketone monitoring in T1DM. Anderson, Lamm, and Hlastala^39^ point to the need for more research on blood-breath associations, keeping in mind that exhaled acetone measurements might underestimate blood levels.

## Conclusion

Breath ketone sensors for diabetes management bear potential to facilitate and improve ketone testing in young people. Our study affirms the key features for young people that drive usability of a breath ketone sensor among this population, and provides a model of user preference assessment among young users. The user experiences with available commercial breath sensors for a ketogenic diet can inform and improve the development of co-designed diabetes-specific breath ketone sensors.

## Data Availability

All data produced in the present study are available upon reasonable request to the authors.

## Abbreviations

DKA: Diabetic Ketoacidosis
T1DM: Type 1 Diabetes Mellitus

## Acknowledgements

This research was funded by and has been delivered in partnership with *Our Health in Our Hands (OHIOH)*, a strategic initiative of the Australian National University, which aims to transform healthcare by developing new personalized health technologies and solutions in collaboration with patients, clinicians, and health care providers. Moreover, this work was supported by a postdoc fellowship of the *German Academic Exchange Service DAAD* (NBS). The authors acknowledge the support of the *Canberra Health Services Paediatric Diabetes Service*. AT gratefully acknowledges the support of the *Australian Research Council* for a *Future Fellowship* (FT200100939) and *Discovery grant* (DP190101864). AT also acknowledges financial support from the *North Atlantic Treaty Organization Science for Peace and Security Programme* project *AMOXES* (#G5634).

## Permission to reproduce material

Not applicable.

## Ethics approval

Ethical approval for the study was obtained by the Australian National University’s Human Ethics Committee and ACT Health Human Research Ethics Committee (HREC, Australia) in April/May 2020 (2019.ETH.00143; 2019/ETH12170). The study participation was based on informed consent (written, signed consent form). For minors, written consent was signed by a parent/guardian.

## Patient consent

The study participation was based on informed consent. All participants signed a consent form.

## Author contribution

All authors have read and approved the final article version. NBS and AP collected the data. CN, KM, and ED advised and guided the sensor selection and sensor testing. NBS and JD analyzed the data, and NBS prepared the manuscript draft. CP contributed to the evaluation design and final review. All authors helped to improve the manuscript. JD, CP, CN, AT, and HS supervised the study.

