## Appendix 1 for "A User Preference Analysis Of Commercial Breath Ketone Sensors to Inform the Development of Portable Breath Ketone Sensors for Diabetes Management in Young People"

**Supplementary material**

**Testing a Breath Ketone Sensor at home – Instructions**

Thank you for agreeing to test a breath ketone sensor at home, we appreciate you taking the time to do this and value your feedback.

The commercially available breath ketone sensor you will be testing is not specifically designed for diabetes. It has been designed for people following a ketogenic diet (see note below). They usually do this for weight loss. We want to know how useful a device like this could be in diabetes management. Your feedback will be the foundation for a breath ketone sensor specifically tailored for diabetes management that we are developing at ANU.

Please read the instructions carefully on how to test the sensor. If you have any questions, don't hesitate to contact the research team (you can find our details below).

1. We will send you a breath ketone sensor, 10 blood ketone test strips and 10 blood glucose test strips. You will need a device to measure blood ketones and blood glucose (for example your FreeStyle Optium Neo). If you do not have one let us know before you start any testing.

2. Set up the breath ketone sensor and practice using it. Read the instructions that come with it and get familiar with how it works. (If the sensor you have received uses an app, download and install the app on your phone, and set it up. Use the app to collect your breath ketone results. Please contact us if you are having difficulty in using it).

3. Once familiar with the breath ketone sensor use, measure your **blood ketones, blood glucose and breath ketones at the same time – two times per day for five days** (10 measurements):

    1. before you have breakfast, and
    2. after school before eating anything

4. Use the **spreadsheet** we have sent you to note down all your measured values (blood ketones, breath ketones, blood glucose, and any others you want to share). Use the comments section to record if you were feeling well or unwell (for example, having a cold or flu), or any other important things we should know.

5. If you would like to do more tests with the breath sensor, you are welcome to do this. Additional breath checks could be:

    1. if your blood glucose is higher than expected
    2. two hours after breakfast
    3. before exercise

4. soon after exercise (before meal)
5. before sleep

6. Again, note down all measurement outcomes in the spreadsheet.

7. Note down **good things about the breath sensor, as well as any problems with it**. You can create a Word document or simply hand write notes on paper.

8. At the end of the testing period, please hand all collected data and information over to us.

**Important NOTE: We do not want you to make any changes to your usual self-management routine. A ketogenic diet is not recommended for people with diabetes, so please just keep everything the way it is...**

<u>**Contact for questions or problems:**</u>

Dr. Nicola Brew-Sam/Dr. Anne Parkinson
Our Health in Our Hands (OHIOH)
Department of Health Services Research and Policy, Australian National University
Mail:
