## Appendix 2 for "A User Preference Analysis Of Commercial Breath Ketone Sensors to Inform the Development of Portable Breath Ketone Sensors for Diabetes Management in Young People"

### Supplementary material

### Interview guide – young people and caregivers

*Instruction: Welcome the participant and inform about audio recording.*

To start with, I would like to ask you a few questions to get some background information:

**1.** How old are you (your birth date)?
**2.** When were you first diagnosed with diabetes (year)?
**3.** Which breath sensor device did you test?
**4.** Can you tell me how often you used the breath sensor during the testing period, and which other measurements you took with other devices.
**5.** Which other technology and devices did you use during the breath ketone sensor testing (CGM, pump, SAPT, etc.; which brands)?
**6.** Were these the same devices you used before the testing started or did you make or have any changes?
**7.** Did you use any phone apps or web apps for diabetes management during the testing period? (If yes, which ones?)
**8.** Do you usually measure your ketone levels? Can you tell me why you measure them and when?

*User experiences:*

**9.** Have you ever used any type of breath sensor, or was this the first time?
   *If previous use:*
   - Which breath sensor(s) did you previously use and why did you use it?
   - What was your experience previously? In particular, why are you still using it/did you stop using it?

   *If no previous use:*
   - Is there any reason why you haven't used a breath sensor before?

*(Un)met expectations and technology characteristics (usability, sensitivity, dynamic range:*

**10.** How easy was it to set up the breath sensor, and did it take long?
**11.** Did the breath ketone sensor work well after you set it up?
**12.** What did you like/dislike about it (features etc.)? *Discuss the following:*
   - Sensor size
   - Sensor shape
   - Battery life
   - Type of result display (traffic light result – red, orange, green – or numerical result)
   - Device design variables (e.g., color)
   - Time to warm up and give results
   - Measurement accuracy
   - Data recording option

- Data sharing option
- Connection to app
- Connection to cloud
- Graphical outputs (e.g., timeline)
- Mouthpieces
- Website company

**13.** What would you change? What would your ideal breath sensor look like?
**14.** Overall, did the sensor meet your expectations and was it useful? Why (not)?
**15.** What do you think are the advantages or disadvantages in using a breath ketone sensor as compared to blood or urine tests?
**16.** Which type of ketone testing do you prefer after having tried the breath sensor (blood, breath, urine testing)? Why?

*Task characteristics (perceived impact):*

**17.** What do you think a breath sensor would change in your daily routine and self-management if you used it long-term (ketone sensor/other breath sensor)?
**18.** If you had a breath sensor long-term, how many times per day would you feel comfortable/willing to measure your ketone levels with the sensor?

*Instruction: Ask for all the data collected within the testing period (spreadsheet, notes, device readings). Thank the participant for their time. Raise awareness to not rely on the commercial breath device if continuing to use it.*
