## Appendix 3 for "A User Preference Analysis Of Commercial Breath Ketone Sensors to Inform the Development of Portable Breath Ketone Sensors for Diabetes Management in Young People"

### Supplementary material

### Interview guide – healthcare professionals

*Introductory question:*

How does it usually work, when and where do patients come see you?

*Ketone measurements:*

- From your experience, when and how do patients check their ketone levels, or get them checked?
- Do you think that regular ketone testing is useful for diabetes management? Why?

*Experiences with breath sensors:*

- Do you have any experiences with breath sensors, and particularly breath ketone sensors? If so, where have you come across these?
- If you have any experiences with breath sensors, could you tell us about your experiences? (e.g., usability, sensitivity, etc.)

*Expectations:*

- What do you think are advantages and disadvantages of breath ketone sensors?
- What would you expect from a breath sensor to be useful for diabetes care?
- Would you expect breath sensors for ketonic diet to be useful for diabetes management?
- Please take a look at the attached pictures of currently available breath sensors for ketonic diet. What do you think needs improvement if these were used for diabetes management?
