## Appendix 4 for "A User Preference Analysis Of Commercial Breath Ketone Sensors to Inform the Development of Portable Breath Ketone Sensors for Diabetes Management in Young People"

### Supplementary material

### Breath and Blood Measurements in the Sample

| IP | Device | Day | Month/ year | Details | Time | Breath ketone (mg/l) | Blood ketone (mmol/l) | Blood glucose (mmol/l) | Unwell | Comments |
|---|---|---|---|---|---|---|---|---|---|---|
| 1 | KetoPro KHC M3 | Day 1 | 06/20 | before breakfast | 08:05 | 0.08 | 0.10 | 6.90 | No | Slight sore throat |
| | | | 06/20 | after school (before eating) | 15:20 | 0.00 | 0.10 | 14.80 | No | |
| | | Day 2 | 06/20 | before breakfast | 08:00 | 0.11 | 0.10 | 10.80 | No | Slight sore throat |
| | | | 06/20 | after school (before eating) | 16:30 | 0.00 | 0.20 | 12.00 | No | |
| | | Day 3 | 06/20 | before breakfast | 08:05 | 0.08 | 0.40 | 9.50 | x | Wondering if the toothpaste used will affect the reading? |
| | | | 06/20 | after school (before eating) | 16:15 | 0.00 | 0.01 | 11.80 | x | |
| | | Day 4 | 06/20 | before breakfast | 08:05 | 0.06 | 0.00 | 9.40 | x | |
| | | | 06/20 | after school (before eating) | 16:30 | 0.00 | 0.01 | 6.20 | x | |
| | | Day 5 | 06/20 | before breakfast | 11:10 | 0.00 | 0.01 | 9.70 | x | Later times today as it's a Saturday |
| | | | 06/20 | after school (before eating) | 16:50 | 0.00 | 0.10 | 8.80 | x | |
| 2 | KetoPro KHC M3 | Day 1 | x | before breakfast | x | 0.66 | 0.20 | x | No | |
| | | | | after school (before eating) | x | 0.66 | 0.10 | x | No | |
| | | Day 2 | x | before breakfast | x | 0.00 | 0.60 | x | Yes | Cold/flu |
| | | | | after school (before eating) | x | 0.00 | 0.20 | x | Yes | Cold/flu |
| | | Day 3 | x | before breakfast | x | 0.00 | 0.00 | x | No | |
| | | | | after school (before eating) | x | 0.00 | 0.00 | x | No | |
| | | Day 4 | x | before breakfast | x | 0.00 | 0.10 | x | No | |
| | | | | after school (before eating) | x | 0.00 | 0.00 | x | No | |
| | | Day 5 | x | before breakfast | x | 0.00 | 0.10 | x | No | |
| | | | | after school (before eating) | x | 0.00 | 0.00 | x | No | |
| 3 | KetoPro KHC M3 | Day 1 | 07/20 | before breakfast | 07:00 | 0.00 | 0.20 | 5.90 | No | |
| | | | | after school (before eating) | 16:00 | 0.00 | 0.00 | 4.80 | No | |
| | | Day 2 | 07/20 | before breakfast | 07:00 | 0.00 | 0.10 | 9.30 | No | |
| | | | | after school (before eating) | 16:00 | 0.00 | 0.10 | 3.60 | No | |
| | | Day 3 | 07/20 | before breakfast | 07:00 | 0.00 | 0.10 | 7.20 | No | |
| | | | | after school (before eating) | 16:00 | 0.00 | 0.10 | 5.90 | No | |
| | | Day 4 | 07/20 | before breakfast | 07:00 | 0.00 | 0.10 | 5.10 | No | |
| | | | | after school (before eating) | 16:00 | 0.00 | 0.20 | 12.10 | No | |
| | | Day 5 | 07/20 | before breakfast | 07:00 | 0.00 | 0.10 | 6.40 | No | |
| | | | | after school (before eating) | 16:00 | 0.00 | 0.00 | 4.90 | No | |

| IP | Device | Day | Month/ year | Details | Time | Breath ketone (mg/l) | Blood ketone (mmol/l) | Blood glucose (mmol/l) | Unwell | Comments |
|---|---|---|---|---|---|---|---|---|---|---|
| | | Day 6 | 07/20 | before breakfast | 07:00 | 0.00 | 0.10 | 4.90 | No | |
| | | | | after school (before eating) | 16:00 | 0.00 | 0.20 | 7.80 | No | |
| | | Day 7 | 07/20 | before breakfast | 07:00 | 0.00 | 0.00 | 6.60 | No | |
| | | | | after school (before eating) | 16:00 | 0.00 | 0.20 | 10.20 | No | |
| | | Day 8 | 08/20 | before breakfast | 07:00 | 0.00 | 0.10 | 5.50 | No | |
| | | | | after school (before eating) | 16:00 | 0.00 | 0.00 | 4.20 | No | |
| | | Day 9 | 08/20 | before breakfast | 07:00 | 0.00 | 0.00 | 6.40 | No | |
| | | | | after school (before eating) | 16:00 | 0.00 | 0.10 | 7.90 | No | |
| 4 | KetoPro KHC M3 | Day 1 | 07/20 | before breakfast | 08:31 | 0.00 | 0.00 | 9.60 | No | |
| | | | | after school (before eating) | 11:16 | 0.01 | 0.10 | 17.40 | No | |
| | | Day 2 | 07/20 | before breakfast | 08:51 | 0.01 | 0.00 | 8.20 | No | |
| | | | | after school (before eating) | 07:33 | 0.00 | 0.00 | 12.70 | No | |
| | | Day 3 | 07/20 | before breakfast | 07:25 | 0.02 | 0.10 | 5.30 | No | |
| | | | | after school (before eating) | 10:26 | 0.01 | 0.10 | 8.30 | No | |
| | | Day 4 | 07/20 | before breakfast | 07:00 | 0.00 | 0.10 | 15.10 | No | |
| | | | | after school (before eating) | 07:25 | 0.00 | 0.00 | 10.40 | No | |
| | | Day 5 | 07/20 | before breakfast | 08:45 | 0.01 | 0.00 | 16.50 | No | |
| | | | | after school (before eating) | 07:37 | 0.00 | 0.00 | 13.60 | No | |

| IP | Device | Day | Month/ year | Details | Time | Breath ketone (g/l) | Blood ketone (mmol/l) | Blood glucose (mmol/l) | Unwell | Comments |
|---|---|---|---|---|---|---|---|---|---|---|
| 5 | House of Keto | Day 1 | 07/20 | before breakfast | 07:41 | 0.00 | 0.00 | 8.40 | No | |
| | | | | after school (before eating) | 15:13 | 0.00 | 0.00 | 7.60 | No | |
| | | Day 2 | 07/20 | before breakfast | 07:25 | 0.00 | 0.10 | 5.10 | No | |
| | | | | after school (before eating) | 15:02 | 0.00 | 0.10 | 3.90 | No | |
| | | Day 3 | 07/20 | before breakfast | 08:03 | 0.00 | 0.10 | 7.60 | No | |
| | | | | after school (before eating) | 15:05 | 0.00 | 0.10 | 4.10 | No | |
| | | Day 4 | 07/20 | before breakfast | 08:19 | 0.00 | 0.10 | 8.30 | No | |
| | | | | after school (before eating) | 14:46 | 0.00 | 0.10 | 7.20 | No | |
| | | Day 5 | 07/20 | before breakfast | 07:08 | 0.00 | 0.10 | 6.60 | No | |
| | | | | after school (before eating) | 14:50 | 0.00 | 0.10 | 9.10 | No | |
| 6 | House of Keto | Day 1 | 07/20 | before breakfast | 07:30 | 0.00 | 0.00 | 12.30 | No | |
| | | | | after school (before eating) | 16:00 | 0.00 | 0.10 | 10.50 | No | |
| | | Day 2 | 07/20 | before breakfast | 07:30 | 0.00 | 0.10 | 12.80 | No | |
| | | | | after school (before eating) | 16:00 | 0.00 | 0.10 | 14.10 | No | |

| IP | Device | Day | Month/ year | Details | Time | Breath ketone (g/l) | Blood ketone (mmol/l) | Blood glucose (mmol/l) | Unwell | Comments |
|---|---|---|---|---|---|---|---|---|---|---|
| | | Day 3 | 07/20 | before breakfast | 07:30 | 0.00 | 0.10 | 6.70 | No | |
| | | | | after school (before eating) | x | x | x | x | x | |
| | | Day 4 | 07/20 | before breakfast | 07:30 | 0.00 | 0.00 | 12.20 | No | |
| | | | | after school (before eating) | 16:00 | 0.00 | 0.10 | 16.40 | | |
| | | Day 5 | 07/20 | before breakfast | 07:30 | 0.00 | 0.00 | 8.60 | No | |
| | | | | after school (before eating) | 16:00 | 0.00 | 0.10 | 13.30 | No | |
| | | Day 6 | 07/20 | before breakfast | 07:30 | 0.00 | 0.10 | 11.30 | No | |
| | | | weekend | before eating | 16:00 | 0.00 | 0.30 | 7.30 | No | I had something to eat and tested 20 minutes after I had forgotten to test first |
| | | Day 7 | 07/20 | before breakfast | 07:30 | 0.00 | 0.10 | 11.20 | No | |
| | | | weekend | before eating | 15:12 | 0.00 | 0.10 | 20.00 | No | High when testing so performed a ketone reading then |
| | | Day 8 | 07/20 | before breakfast | 07:30 | No reading | No reading | 7.30 | No | |
| | | | | after school (before eating) | 16:00 | 0.00 | 0.10 | 13.50 | No | |
| | | Day 9 | 07/20 | before breakfast | 07:30 | 0.00 | 0.00 | 5.10 | No | |
| | | | | after school (before eating) | 16:00 | 0.00 | 0.10 | 12.70 | No | |
| 7 | House of Keto | Day 1 | 08/20 | before breakfast | 08:20 | 0.00 | 0.10 | 11.00 | No | Generally well |
| | | | | after school (before eating) | 16:00 | 0.00 | 0.10 | 7.10 | No | Generally well |
| | | Day 2 | 08/20 | before breakfast | 09:00 | 0.00 | 0.10 | 6.90 | No | Generally well |
| | | | | after school (before eating) | 17:00 | 0.00 | 0.10 | 15.00 | No | Generally well |
| | | Day 3 | 08/20 | before breakfast | 07:00 | 0.00 | 0.10 | 5.70 | No | Generally well |
| | | | | after school (before eating) | 16:30 | 0.00 | 0.10 | 14.10 | No | Generally well |
| | | Day 4 | 08/20 | before breakfast | 07:15 | 0.00 | 0.10 | 13.00 | No | Generally well |
| | | | | after school (before eating) | 16:45 | 0.00 | 0.10 | 2.80 | No | Generally well |
| | | Day 5 | 08/20 | before breakfast | 07:00 | 0.20 | 0.10 | 11.60 | No | Well |
| | | | | after school (before eating) | 18:00 | 0.00 | 0.10 | 5.20 | No | Well |
| 8 | House of Keto | Day 1 | x | before breakfast | 08:58 | 0.00 | 0.00 | 4.00 | No | |
| | | | | after school (before eating) | 18:07 | 0.01 | 0.10 | 8.60 | No | |
| | | Day 2 | x | before breakfast | 09:28 | 0.00 | 0.30 | 7.70 | No | |
| | | | | after school (before eating) | 18:17 | 0.00 | 0.10 | 7.80 | No | |
| | | Day 3 | x | before breakfast | 07:55 | 0.00 | 0.10 | 7.10 | No | |
| | | | | after school (before eating) | 18:16 | 0.00 | 0.10 | 5.70 | No | |
| | | Day 4 | x | before breakfast | 07:45 | 0.00 | 0.10 | 8.80 | No | |
| | | | | after school (before eating) | x | x | x | x | x | |
| | | Day 5 | x | before breakfast | x | x | x | x | x | |
| | | | | after school (before eating) | x | x | x | x | x | |
| 9 | House of Keto | Day 1 | 08/20 | before breakfast | 07:00 | 0.00 | 0.10 | 12.00 | x | |

| IP | Device | Day | Month/ year | Details | Time | Breath ketone (g/l) | Blood ketone (mmol/l) | Blood glucose (mmol/l) | Unwell | Comments |
|---|---|---|---|---|---|---|---|---|---|---|
| | | | | after school (before eating) | 16:30 | 0.02 | 0.30 | 18.60 | x | No sensor - no lunch bolus |
| | | Day 2 | 08/20 | before breakfast | 08:30 | 0.00 | 0.10 | 15.60 | x | BGL 15.1 |
| | | | | after school (before eating) | x | x | x | x | x | |
| | | Day 3 | 08/20 | before breakfast | 08:30 | 0.00 | 0.10 | 7.40 | x | |
| | | | | after school (before eating) | 15:30 | 0.00 | 0.10 | 8.80 | x | BGL 9.5 - 8.6 |
| | | Day 4 | 08/20 | before breakfast | 07:00 | 0.00 | 0.10 | 9.30 | x | BGL 10.2 |
| | | | | after school (before eating) | x | x | x | x | x | |
| | | Day 5 | 08/20 | before breakfast | 07:00 | 0.00 | 0.10 | 7.80 | x | BGL 7.5 |
| | | | | after school (before eating) | 15:31 | 0.00 | 0.00 | 4.80 | x | |
| | | Day 6 | 08/20 | before breakfast | 07:00 | 0.00 | 0.10 | 5.80 | x | BGL 6.6 |
| | | | | after school (before eating) | 16:30 | 0.00 | 0.10 | 7.40 | x | BGL 7.7 |
| | | Day 7 | 08/20 | before breakfast | 07:00 | 0.00 | 0.00 | x | x | |
| | | | | after school (before eating) | x | x | x | x | x | |
| | | Day 8 | 08/20 | before breakfast | 07:00 | 0.00 | 0.00 | 8.10 | x | BGL 8.8 |
| | | | | after school (before eating) | x | x | x | x | x | |
| | | Day 9 | x | before breakfast | 08:15 | 0.00 | 0.00 | 7.80 | x | |
| | | | | after school (before eating) | 15:15 | 0.00 | 0.10 | 8.60 | x | |
| | | Day 10 | x | before breakfast | 09:30 | 0.00 | 0.10 | 12.70 | x | BGL 14.4 |
| | | | | after school (before eating) | x | x | x | x | x | |
| 10 | House of Keto | Day 1 | 08/20 | before breakfast | 08:15 | 0.00 | 0.00 | 7.60 | No | |
| | | | | after school (before eating) | 02:15 | 0.00 | 0.00 | 5.70 | No | |
| | | Day 2 | 08/20 | before breakfast | 11:50 | 0.00 | 0.00 | 6.10 | No | |
| | | | | after school (before eating) | 02:15 | 0.00 | 0.00 | 4.20 | No | |
| | | Day 3 | 08/20 | before breakfast | 08:10 | 0.00 | 0.00 | 7.80 | No | |
| | | | | after school (before eating) | 19:20 | 0.00 | 0.00 | 9.50 | No | |
| | | Day 4 | 08/20 | before breakfast | 15:40 | 0.00 | 0.00 | 8.20 | No | |
| | | | | after school (before eating) | 22:00 | 0.00 | x | 7.80 | No | |
| | | Day 5 | 08/20 | before breakfast | 16:00 | 0.00 | 0.10 | 12.00 | No | |
| | | | | after school (before eating) | 22:00 | 0.00 | 0.10 | 10.50 | No | |
| | | Day 6 | 08/20 | night | 02:00 | 0.00 | 0.20 | 17.00 | No | Just a random test because of high BGL |

*Notes*. BGL= blood glucose level
